# Genome sequencing combining prenatal ultrasound in the evaluation of fetal CNS structural anomalies

**DOI:** 10.1101/2020.03.04.20031294

**Authors:** Ying Yang, Sheng Zhao, Guoqiang Sun, Fang Chen, Tongda Zhang, Jieping Song, Wenzhong Yang, Lin Wang, Nianji Zhan, Xiaohong Yang, Xia Zhu, Bin Rao, Zhenzhen Yin, Jing Zhou, Haisheng Yan, Yushan Huang, Jingyu Ye, Hui Huang, Chen Cheng, Shida Zhu, Jian Guo, Xun Xu, Xinlin Chen

**Author notes:** Contributed equally as corresponding authors Corresponding author contact information (Xinlin Chen). Contributed equally as first authors (Ying Yang, Sheng Zhao, Guoqiang Sun).

## Abstract

**Purpose:** Genome sequencing (GS) is potentially the most suitable diagnostic tools for fetal CNS structural anomalies. However, its efficacy hasn’t been proved in large cohort of fetal CNS structural anomalies.

**Methods:** Patients were enrolled by a multiple-level referral system when fetal CNS structure anomalies were found by ultrasonography. Samples from fetuses were subjected to GS.

**Results:** Data of 162 fetuses with 11 frequent types of CNS anomalies was collected. The overall diagnosis yield of GS was 38.9%. 36(20.3%) fetuses were detected with chromosomal anomalies and pathogenic CNVs. Pathogenic or likely pathogenic single-gene variants and intragenic CNVs were found in 24 and three fetuses, contributing 14.8% and 1.9% diagnostic yield respectively. The diagnostic rate in 41 fetuses with CNS malformation combined with anomalies out of brain was as high as 73.3%. Malformations of the posterior cerebral fossa, abnormal neuronal proliferation and migration have the highest diagnostic rates. NTDs had the second lowest diagnostic rates of 14.7% and none pathogenic variants were found in ultrasound anomalies that suggested destructive cerebral lesions.

**Conclusion:** GS is an efficient genetic testing tool with the diagnostic power compared to current CMA plus ES procedure in fetal CNS anomalies evaluation.

## Introduction

Fetal central nervous system (CNS) anomalies are major indications for termination of pregnancy and there are high possibilities of causing severe consequences such as intrauterine and infant deaths and moderate or severe disabilities.^1^ Screening and diagnosing CNS anomalies correctly as early as possible to provide more information for clinical decisions is important.

Ultrasonography is the most important modality for evaluation of fetal growth and anatomy with high efficiency and specificity of detecting congenital anomalies prenatally, especially in CNS.^2^ Fetal MRI is a second image modality that adds useful information in fetal CNS evaluation. ^3^ It is generally reported that around 80%-90% of fetal CNS malformations can be diagnosed correctly.^3^

Genomic variations are important etiologies of fetal anomalies. Karyotyping and chromosomal microarray analysis (CMA) are effective in diagnosing fetal malformations caused by chromosomal anomalies and copy number variations (CNVs).^4^ Eome sequencing (ES) is usually applied after uninformative results of CMA in currently clinical practice to detect single-gene mutations on exomes.^5,6^ This strategy may be economical but it overlooks the fact that pathogenic genomic variation varies in different congenital diseases. For fetuses, time is most essential. Therefore, a one-for-all genetic testing modality is necessary.^7^ It has been shown that low-pass genome sequencing (GS) provided additional information in prenatal setting as compared with CMA.^8^ High-depth GS has been reported to expand diagnostic utility in infants or children with a wide spectrum of disorders.^9,10^ However, the diagnostic potential of GS hasn’t been evaluated in large cohort of fetal ultrasound anomalies. Since the cost of GS is decreasing rapidly, it is not necessarily more expensive than the cost of CMA and ES together. Therefore, it is both necessary and possible to investigate the diagnostic and research utilities of GS in large-scale studies of fetal anomalies.

Here, we displayed the GS data of 162 unselected fetal ultrasonography defined CNS anomalies form our large cohort of fetal anomalies to evaluate the diagnostic power of GS in comparison of current popular genetic testing modalities in fetal CNS ultrasound anomalies. Besides, this study also demonstrated the distribution of a full spectrum of genomic variations and their characteristics in major types of ultrasound fetal CNS anomalies including multifactorial diseases such as neural tube defects (NTDs) and hydrocephalus for more compressive and better understanding of CNS anomalies.

## Method

### Patient identification, ultrasound examination and sample collection

Pregnant women who were found fetal structural malformation by routine prenatal ultrasound screening in a multiple-level referral system of prenatal screening and diagnosis were recruited for a large-scale project for studying the genomic variations of all kinds of congenital defects. This referral system was launched and conducted by Maternal and Child Health Hospital of Hubei Province (HBFY), which includes 156 prenatal screening units and 32 diagnostic centers all over Hubei Provence as well as several districts in middle and southwestern China. All of the screening units and diagnostic centers were trained by HBFY followed ISUOG practice guideline.^11^ Then patients were transferred to HBFY for thorough systematic or special prenatal ultrasound by at least two chief physicians to get a final diagnosis. MRI was also provided when it was accepted by patients and their families. Altogether, 162 patients with fetal CNS anomalies agreed to donate the fetal samples (161 of umbilical cord tissue samples after birth or termination of pregnancy, and one amniotic fluid sample) with signed informed consents. Demographic information has been asked from the pregnant women but they could refuse to answer. Pregnancy outcomes were independent of genetic diagnosis of this study. This project followed Chinese state and local regulations related to biological and medical researches and was approved by HBFY Ethics Committee.

### Genome sequencing

Genomic DNA from fetal tissues were extracted using salting out method as previously described by Miller S. et al.^12^ and examined by Qubit 3.0 fluorometer (LifeTechnologies, Paisley,UK) and 1% agarose gel electrophoresis. Sequencing library was constructed and sequenced according to manufacturer’s instructions for the BGISEQ500 or MGISEQ2000 sequencer platform.^13^ The read length was pair-end 100bp.

### Variants calling, annotation and interpretation

Chromosomal anomalies including aneuploidy and rare copy number variation were called based on the pipeline developed by Dong et al^14^. Intragenic CNVs that smaller than 100Kb were called by SpeedSeq SV pipeline (v.0.0.3a).^15^ Deep sequencing data were aligned to the human reference genome GRCh37/hg19 and single-nucleoside variants(SNV, including insertion/deletion) were called and filtered using the Edico Genome’s Dragen Bio-IT Platform.^16^ Intragenic CNVs and SNVs were annotated using bcfanno(v1.4) (https://github.com/shiquan/bcfanno) with frequencies in public database (ExAC, GnomAD and G1000^17^), in-house normal population database containing 790 samples sequenced by the same sequencing platform. Variants with MAF higher than 0.01 in above mentioned databases were filtered.^18^

Clinical significance of variants was determined by a clinical review panel that consists of genetics, senior sonographers, MRI radiologist and obstetricians and gynecologists. Variants were interpreted based on the American College of Medical Genetics and Genomics (ACMG) guidelines^19,20^ to pathogenic/likely pathogenic or of unknown significance (VUS). Besides we defined an extra class of candidate variants that on genes known to associated with CNS anomalies and had not presented in public database or inner database if the gene is associated with dominant diseases, or two variants on a same gene associated with recessive inheritance diseases.

### Validation

Pathogenic or likely pathogenic SNV were validated by Sanger sequencing. CNVs of duplication were validated by QPCR and CNVs of deletions were validated by PCR and agarose gel electrophoresis.

### Data acquirement

The data reported in this study are available in the CNGB Nucleotide Sequence Archive (CNSA: CNP0000415).

## Results

### Demographic characteristics of the cohort and prenatal imaging features

The median maternal age is 27. No difference was detected in the age median and distribution between this cohort and that of the Chinese women who gave a birth in 2015 calculated from the sample survey data by National Bureau of Statistics of China. ^21^ The chromosomal gender ratio of the fetuses is 1:1(51/51), suggesting no gender difference of this cohort. Above 60% of the final ultrasound diagnosis were got after 28weeks (Supplementary Figure S1).

Clinical information and detailed prenatal ultrasound or MRI features of each individual was reviewed and listed in supplementary Table S1. CNS anomalies were divided into 12 subgroups and their co-existing connections were shown in Figure. 1c. Hydrocephalus and NTDs were most frequent in our cohort. 117 cases found anomalies in the CNS only. The rest 45 ones were combined with abnormalities out of CNS.

**Figure 1.**
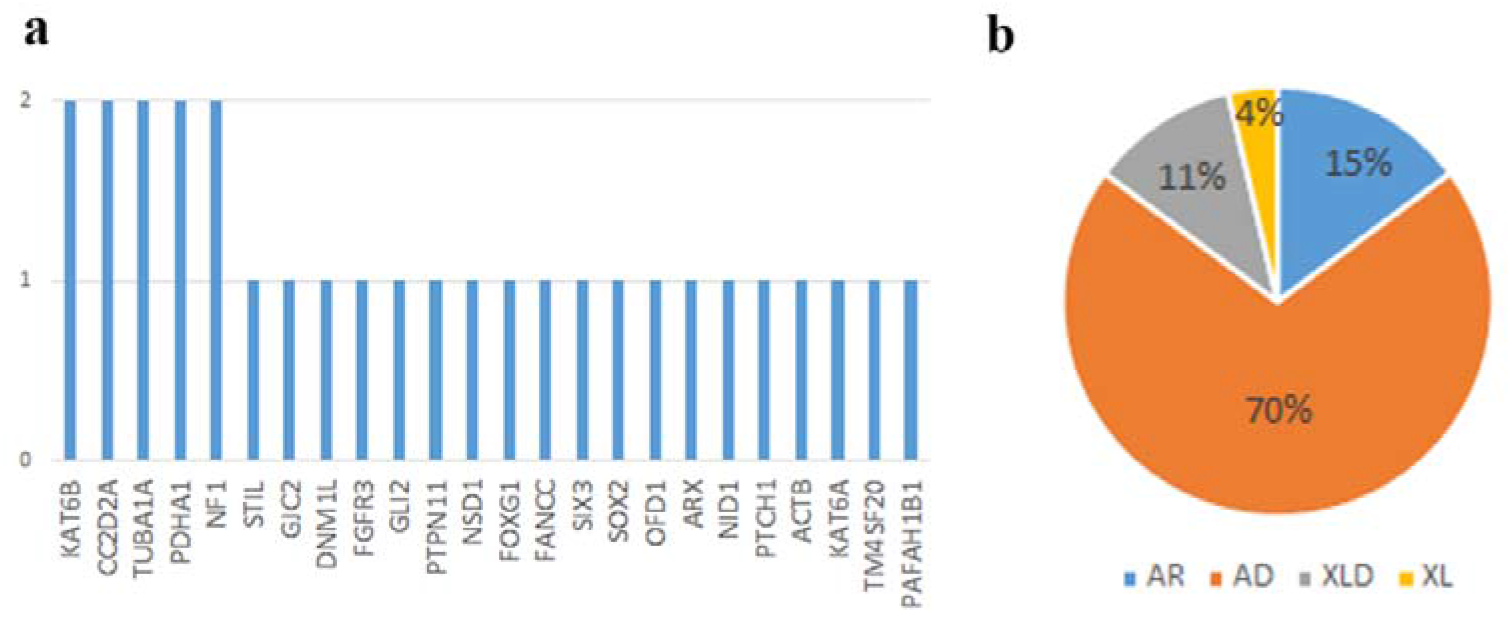
Critical genes identified in diagnosed cases and inheritance patterns. a). Frequency of genes with diagnostic variants or critical genes in pCNVs. b). Percentage of inheritance modes of diseases associated with genes in a), AR, autosome recessive; AD, autosome dominant; XLD, X-linked dominant.

### Chromosomal anomalies and CNVs

Chromosomal anomalies referred to aneuploidies and chromosomal rearrangements over 5Mb to compare with a 550-band resolution karyotype. CNVs referred to those >100Kb that is the same discrimination level as most commercial CMA products. 33(20.3%) fetuses were detected with chromosomal anomalies, including 19 aneuploidies and 14 other chromosomal anomalies and 336 CNVs (Figure S2) containing six pathogenic or likely pathogenic ones (pCNVs) shown by Table 1. In three fetuses, we detected both chromosomal anomalies and pCNVs, implicating imbalanced translocations. Altogether, 36 fetuses got a genetic diagnosis, making a diagnosis yield of 22.2% at the level of 100Kb and above.

**Table 1.** Chromosomal anomalies and pathogenic CNVs.

| Patient No. | pathogenic/likely pathogenic CNV | Size | Critical genes |
| --- | --- | --- | --- |
| 79, 102, 240, 103, 350, 482 | +18 |  | NA# |
| 155, 351, 438, 871, 747, 901 | +13 |  | NA |
| 827, 1008, 1142, 1506 | +21 |  | NA |
| 112, 772 | 45XO |  | NA |
| 1133 | 47XXY |  | NA |
| 938 | del(1p36.2p36.3).seq(823534-15632453)x1 | 14.8Mb | NA |
| 864 | dup(2q36.3q37.3).seq(226537458-242997727)x3<br>del(6q26q27).seq(161507517-170879606)x1 | 16.5Mb<br>9.4Mb | NA |
| 470 | del(3q12.1q21.2).seq(99540858-125130561)x1 | 25.5Mb | NA |
| 244 | del(4q31.3q32.1).seq(153436540-160087839)x1 | 6.6Mb | NA |
| 186 | dup(5p14.3p15.3).seq(10429-23273021)x3<br>del(18p11.3p11.3).seq(111935-4272634)x1 | 23.2Mb<br>4.2Mb | NA |
| 116 | del(6q25.3q27).seq(160630268-170879606)x1 | 10.2Mb | NA |
| 456 | dup(7p22.1).seq(5029498-6809995) x3 | 1.8Mb | ACTB |
| 190 | del(7q35q36.3).seq(149299056-159068966)x1 | 9.8Mb | NA |
| 238 | del(7q33q36.3).seq(137529688-159068966)x1 | 2.2Mb | NA |
| 422 | del(7q35q36.1).seq(145049787-159068966)x1<br>dup(19q13.4).seq(55330867-59044235)x3 | 14.0Mb<br>3.7Mb | NA |
| 653 | del(8p11.21).seq(41835654-41836946)x1 | 1.3Kb | KAT6A(EX6) |
| 532 | dup(9p24.1p24.3).seq(10001-6476812)x3 | 6.5Mb | NA |
| 664 | del(13q22.1q34).seq(74369596-115054392)x1 | 40.7Mb | NA |
| 931 | dup(13q31.2q34).seq(89863557-115054392)x3<br>del(20p13).seq(60001-2226344)x1 | 25.2Mb<br>2.2Mb | NA |
| 796 | del(17p13.2p13.3).seq(1081133-4774754) x1 | 3.7Mb | PAFAH1B1 |
| 882 | dup(17p13.3).seq(1150479-1592862) x3 | 442Kb | BHLHA9 |
| 884 | del(17q11.2).seq(29447084-29503935)x1 | 56.8Kb | NF1(EX2-EX5) |
| 954 | del(17q11.2).seq(228230706-228234866)x1* | 4.16Kb | TM4SF20(EX3) |
| 810 | del(18p11.2p11.3).seq(111935-15323954)x1 | 15.2Mb | NA |
| 1133 | del(18p11.2p11.3).seq(111935-15334797)x1 | 15.2Mb | NA |

#: NA: not applicable
\*: with an ~100bp gap within it

### Single nucleotide variants and Intragenic CNVs

Except 26 samples that have been found chromosomal anomalies at low-passage, 136 samples were deeply sequenced to an average depth of 41.9± 0.4-fold coverage. Details of sequencing data and variant calling results were seen in Table S2.

26 pathogenic or likely pathogenic SNVs in 20 genes that may contribute to the fetal CNS malformations were identified in 24 cases, as shown in Table 2. Among them, 15 ones are novel mutations. Seven ones are previously known pathogenic variants and the rest four are found in public databases but without clear clinical significance before.

**Table 2.**
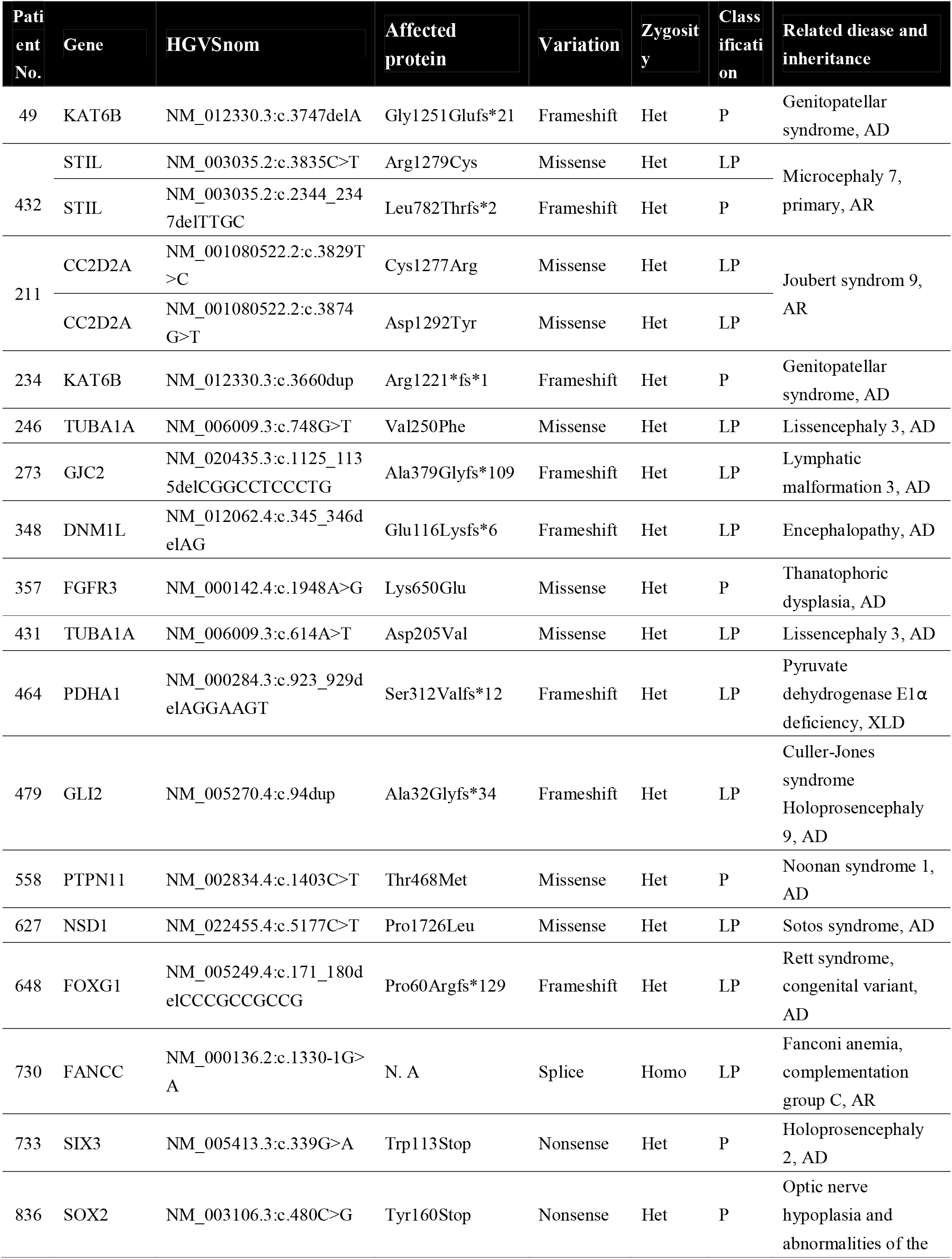
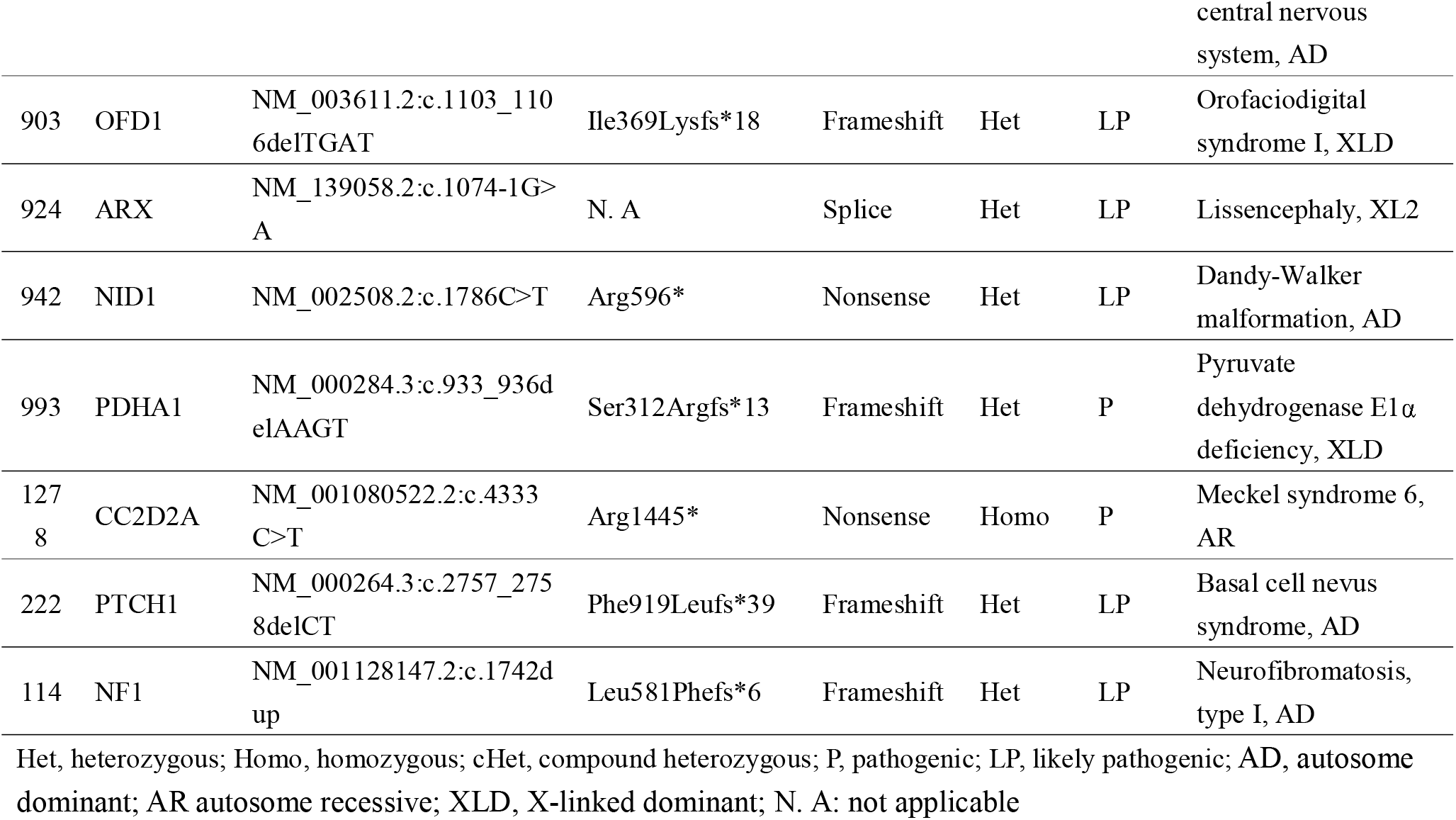
Pathogenic or likely pathogenic SNVs identified.

We also found 24 candidate SNVs in 22 cases (Table S3,Figure 3) might be related with the anomalies as defined in Methods. Knowing origins of these mutations would add extra information in determination their clinical significance but unfortunately almost all samples are lacking of parental DNA in this study. Besides, there are three secondary findings and four incident findings.

Intragenic CNVs ranging from 50bp to 100Kb that beyond the detection limit of mainstream CMA were analyzed in 102 samples without chromosomal anomalies, pCNVs or pSNVs. Four samples didn’t pass the quality control test due to lower quality of extracted genomic DNA. On average 2866.2±40.9 intragenic CNVs were called in each sample as shown by Figure 2a. The number of CNVs containing genes are linearly dependent on the total numbers of CNVs (Figure 2b). Numbers of exon-containing CNVs seemed less but also related with the number of CNVs called in the sample.

**Figure 2.**
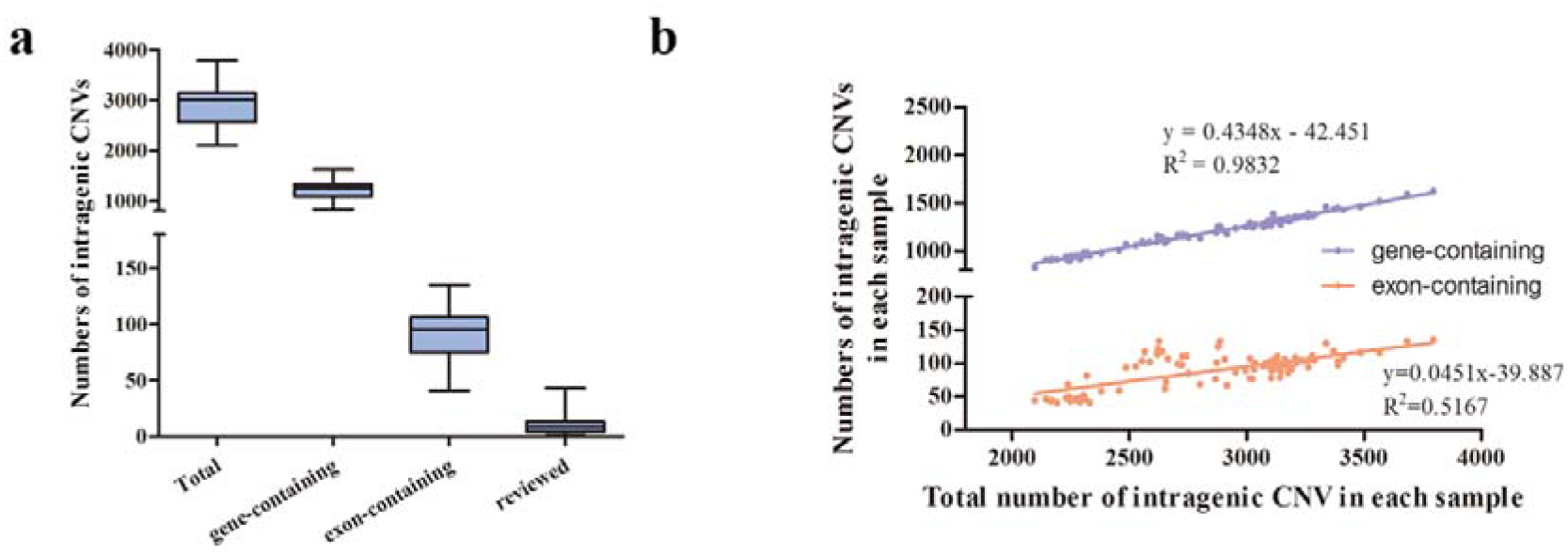
GS’s power of identifying intragenic CNVs. a). Numbers of total intragenic CNVs, gene-containing, exon-containing and panel-reviewed CNVs. b) Linear relationship between the number of total intragenic CNVs and that of CNVs containing genes and that of exon-containing CNVs.

Classification rules for CNVs and SNVs were both applied for intragenic CNVs. Only CNV containing exons and with a frequency lower than 0.01 in our in-house database were reviewed by clinical review panel. As a result, for each sample there was on average 10.8 variants assessed. We found likely pathogenic intragenic CNVs that may associate with the sonographic abnormalities in three cases (Table 1). In P653 found bilateral ventriculomegaly, subependymal cysts and development delay, a 1292bp deletion on exon6 of KAT6A was identified. We didn’t find any similar deletions in DGV or previous publications, but protein truncating mutation such as frameshift and nonsense mutations of KAT6A is an established genetic cause for autosomal dominant mental retardation characterized by microcephaly and global developmental delay with intellectual disability. ^22^ Tham et al ^23^ reported MRI changes such as altered anterior horn of lateral ventricles and cystic periventricular leukomalacia. In P884, a 56.8Kb deletion (17: 29447084-29503935) on NF1 expanding exon2-exon5 was identified. This deletion hasn’t been reported before to our knowledge, but there is a pathogenic CNV (17:29425306-29488097) in Decipher overlapping more than 70% of what we found. Besides, single or multi-exons deletions expanding exon2-4 have been found in NF1 patients.^24^ We found hypoplasia of corpus callosum, enlarged third ventricle and small cysts on cerebral midline by prenatal ultrasound in P884, which may corresponded to frequently observed structural abnormalities of the brain by MRI in NF1 children such as enlargement of the corpus callosum^25^, abnormalities of white matter microstructure in frontal lobes and corpus callosum^26^ and hyperintensity lesions in T2 MRI^27^, suggesting that prenatal sonographic features of NF1 might be related with postnatal imaging phenotypes. A third likely pathogenic intragenic CNV was a complex 4160bp deletion (2: 228230706- 228234866) consisting of 2 microdeletions with a gap of 100bp in P954. Prenatal sonography of the fetus showed widen left lateral ventricle and abnormal hypoechoic signals around bilateral anterior horns. This intragenic deletion contained exon3 of TM4SF20 gene and has been described by Wiszniewski W et al ^28^ which is ancestral deletion and segregates with early language delay disorders and cerebral white matter hyperintensities (WMH) in 15 unrelated families predominantly from Southeast Asian population. In the study one of the severely affected patient who was prematurely born at 33-week gestation presented gross enlargement of the third and lateral ventricles with near-complete loss of periventricular white matter, while other patients showed relatively milder but varying degrees of WMH.^28^

Among these causative genes including critical genes in pCNV, there are five recurrent ones: TUBA1A, KAT6B, CC2D2A, and PDHA1 and NF1 seen in Figure 1a. About 70% of them are related with autosome dominant diseases and 15% with autosome recessive inheritance. The rest ones are X-linked or X-linked dominant, as shown by Figure 1b. The genes are largely connected in regard of protein-protein interaction with a significant p-value (1.75e-10) according to STRING database(http://string-db.org) network interaction analysis and significantly enriched in brain development, forebrain development and anatomical structure morphogenesis (Figure S3).

Compound heterozygous of SNV and intragenic CNV were not found in this study. In total, SNVs analysis brought a diagnostic yield of 14.8% in the whole study cohort of 162 patients, and an extra diagnostic rate of 19.0% based on uninformative CNVs Analysis of intragenic CNVs brought an extra diagnostic yield of 2.9% (3/102) on the uninformative CNVs and SNVs, consisting 1.8% (3/162) to the total diagnostic rate of the cohort.

### The potential of combining prenatal imaging and genomic sequencing both in clinic and in research

The diagnostic yield of GS varies significantly among different ultrasound well-defined CNS anomalies with an overall 38.9% (63/162) diagnosis yield. As shown in Figure 2, malformations of the posterior cerebral fossa such as hypoplasia of cerebellum or cerebellar vermis and Dandy-Walker variants, malformations of midline structures such as holoprosencephaly and aplasia/hypoplasia of corpus callosum are mostly related with causative genome variants followed by intracranial cyst, ventriculomegaly and microcephaly. As shown by Figure 3, our diagnostic rate in brain anomalies companied with other organs is over 70% while it is about 30% in cases with only CNS malformations. Prenatal imaging features are extreme useful to efficiently target candidate genes and classify variants and thus are essential to the final diagnosis of GS.

**Figure 3.**
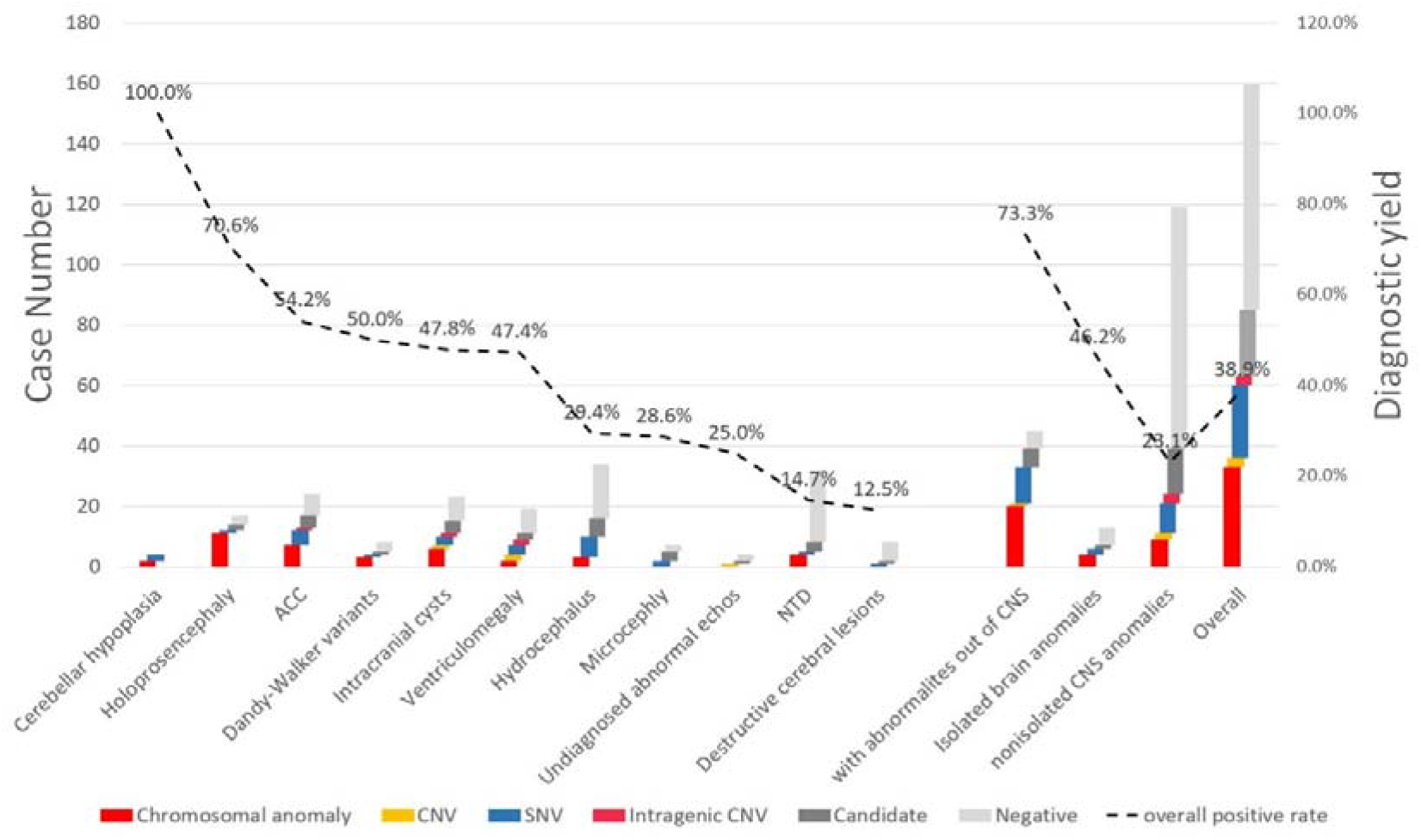
Diagnostic rate of GS and diagnostic variant type distribution. Different diagnostic yields in each sonographic CNS abnormities and subgroups of CNS anomalies only and multiple anomalies out of CNS were demonstrated. Bars indicated case numbers and the dashed line showed the diagnostic rate with the percentages on it. Cases are repeatedly counted if they have more than one sonographic feature. Isolated brain anomalies included only aplasia of corpus collasum, arachnoid cyst and ventriculomegaly as other CNS anomalies usually are complex and involves several anatomic structures of the brain. ACC: aplasia of corpus collasum.

Hydrocephalus and NTDs, the two largest subgroups, were found lower diagnosis rate as it is well known that they have complex etiologies including genetic, environmental and multiple factors^29,30^. The diagnostic rate in hydrocephalus is relatively lower than other types of CNS anomalies, however it is already higher than what has been reported ever before^31^ and more than 2/3 of diagnosed cases were found with pSNVs. Protein truncating mutations in PDHA1 were recurrent found in P464 and P993, suggesting pyruvate dehydrogenase E1-alpha deficiency was one of the important causes of fetal hydrocephaly in the third trimester. Besides, Fanconi anemia is a second major cause of fetal hydrocephaly since we found a pathogenic homozygous splicesite mutation in FANCC in P730 and candidate compound heterozygous mutations in ERCC4 known for related with Fanconi anemia in P54.

What worth mentioning that we identified seven pathogenic or likely pathogenic variants in eight cases with NTDs. Four of them are chromosomal anomalies (seen in Table 1), three SNVs and 1 intragenic CNV. A homozygous nonsense mutation c.4333C>T(p.Arg1445*) in CC2D2A causing Meckel Grubel syndrome was confirmed in P1279. In addition, we found a known pathogenic missense mutation c.1744G>A(p.Gly582Ser) in COL3A1 reported to cause Ehlers-Danlos syndrome IV^32^ (EDS) in P175 with spina bifida occulta at sacral vertebrae. In a second NTD case, P866 diagnosed with exencephaly at 14 week, a heterozygous missense variant with unknown significance on COL3A1 c.542C>T(p.Pro181Leu) was identified. Allele frequency of this variant in ExAC_EAS is 0.0002 and multiple lines of computational evidence support a deleterious effect. Moreover, we found a canonical splice site mutation c.2032+1G>A in ELN in P37 with meningocele (Table S3). ELN has been reported to be causative to autosome dominant cutis laxa. COL3A1 and ELN are both related with skin pathologies and it’s well known that collagen and elastin are major structural components of extracellular matrix^33^. Although there are no previous studies indicating relations of COL3A1 and ELN with NTD, a study of EDS reported 11 patients found with COL3A1 variants and one showed spina bifida, hydrocephalus and hypermobility^34^. Though complex causal genetics of NTD still largely remained unclear, our findings may suggest a role of skin pathology in NTD.

On the other hand, sequencing results are of great significance in helping clarifying sonographic diagnosis. For example, P211 were found ventriculomegaly, hyperechoic lesions in periventricular zone, and cardiac space-occupying lesions (suspecting rhabdomyoma) accompanied with pericardial effusion and pleural effusion seen in Figure 4a-d. It was suspected to be tuberous sclerosis (TS) but ultrasound and MRI manifestation of the intracranial lesions seemed atypical. GS identified none potential damage variants in TSC1 and TSC2, instead a frameshift mutation NM_000264.3:c.2757_2758delCT (p.Phe919Leufs*39) in PTCH1 was found, suggesting the basal cell nevus syndrome rather than TS. It’s characterized by multiple nevoid basal-cell epitheliomas and broad phenotypes including cardiac fibroma and intracranial calcification. Another case was P882 with a 1.7*1.3cm well-defined hypoechoic lesions at the right side of cerebral midline, and widths of right lateral ventricle and cisterna magna were slightly over upper limits. We detected a 440Kb microduplication on 17p13.3(17:1150479-1592862) in it, and similar CNVs was reported previously to related with developmental delay and autism as well as hypoplasia of cerebellum and corpus collasum^35^. 17p13.3 is a critical region for brain cortical morphology. Besides the microduplication, we also identified in P796 a 3.7M microdeletion (17:1081133-4774754) covering critical genes PAFAH1B1 of Miller-Dieker syndrome also known as 17p13.3 deletion syndrome which is characterized by lissencephaly and craniofacial dysmorphism^35,36^. This fetus was found the width of right cerebral lateral ventricle to be 1.04cm by sonography and then MRI revealed abnormal cortical gyration and autopsy confirmed the smooth brain cortex (Figure 4e-h). Therefore, GS added significant useful information in situations that were difficult to get a clear prenatal diagnosis.

**Figure 4.**
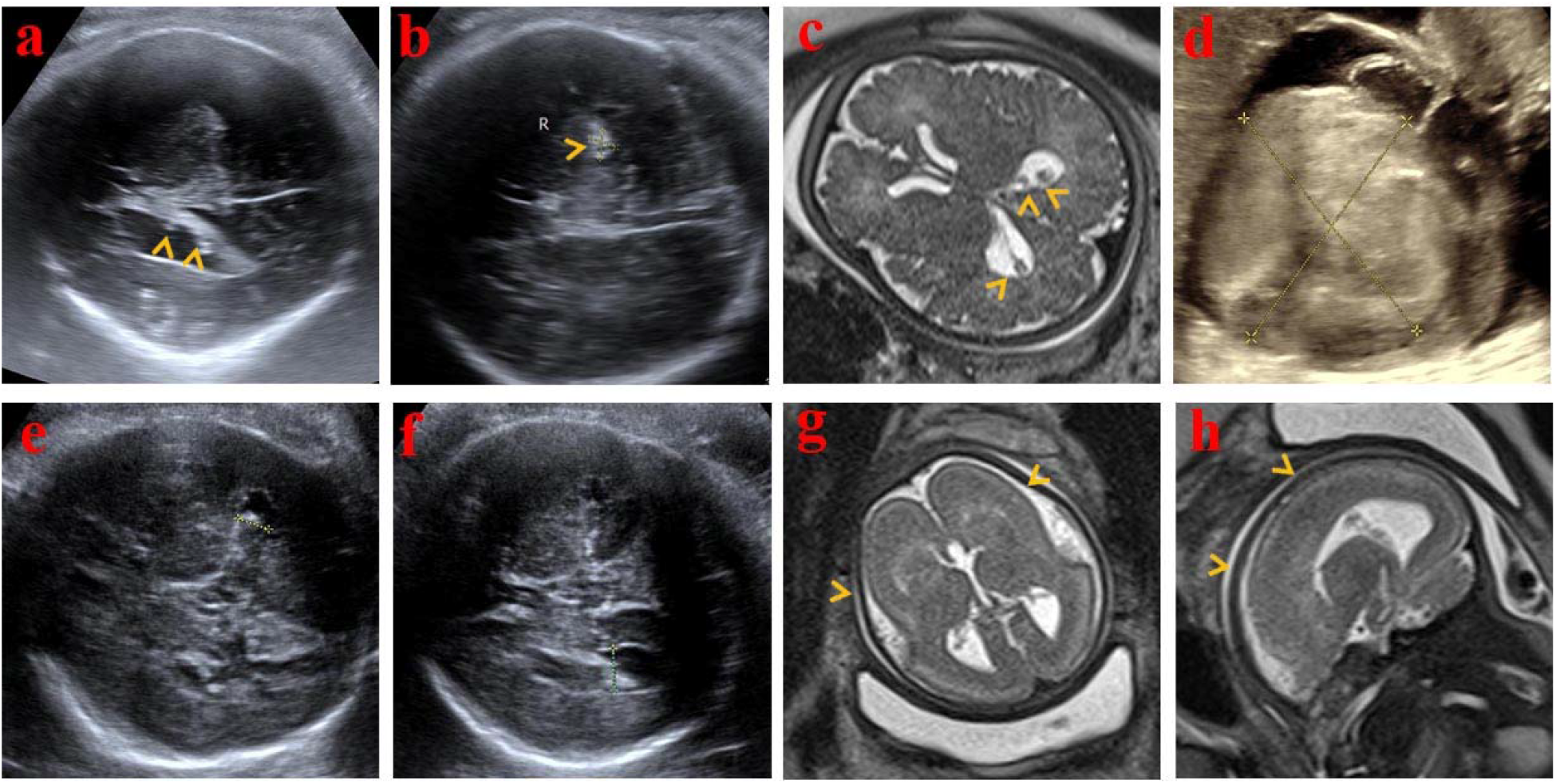
Imaging of two typical cases, P221 (top row) and P796 (bottom row). Sonographic hyperechoic lesions in a) cerebral lateral ventricles and b) brain parenchyma indicated by yellow arrow heads. c) Hypointensity lesions revealed by T2-weighted MRI in cerebral lateral ventricles as pointed out by yellow arrow heads. d) Huge space-occupying lesion in left ventricle shown by d) sonography. Widths of cerebral ventricle of P796 were 0.84cm on the left e) and 1.04cm on the right f) shown by sonography. g) coronal and h) sagittal view of T2 MRI showing abnormal cortical gyration indicated by yellow arrow heads.

## Discussion

We got a final 38.9% (63/162) diagnostic performance of GS in this cohort, which is higher than any other single genetic testing tool currently used in clinic including karyotype, CMA, ES in fetuses with structural malformations^4-6,37^. As indicated in Table 3, diagnostic yields of genomic variants at different resolution level reported by previous studies with large sample size were compared. At microscopic level, our diagnostic rate was close to karyotyping except that balanced translocation and triploid analysis was not applicable by our GS approach. The diagnostic yield of submicroscopic level was lower than what was previously reported. There could be several reasons. First and most important, we have a high proportion of NTD and hydrocephalus cases and small proportion of posterior fossa defects while NTD and hydrocephalus have the lowest diagnostic yield and posterior fossa defect has highest diagnostic rates, which is consistent of Shaffer’s result^38^ by microarray. Besides, these studies usually applied CMA on the basis of normal karyotyping, but the resolution of karyotyping may vary from 5 to10Mb between studies. In Shaffer’s study, the resolution of karyotyping was 10Mb, and it’s not mentioned in Fu’s study.

**Table 3.** Comparison of diagnostic yields for variants at different resolution level.

| Variants resolution | Diagnostic yield of publications (Significant/total cases) |  |  |  |  |  |
| --- | --- | --- | --- | --- | --- | --- |
|  | Publications |  |  | Our study |  |  |
|  | CNS only | CNS + other | CNS total | CNS only | CNS + other | CNS total |
| Microscopic level |  |  |  |  |  |  |
| Chromosomal anomaly | 8.5% (77/908)<sup>37</sup> | N. A | 18.4% (37/201)<sup>42</sup> | 11.1% (13/117) | 44.4% (20/45) | 20.4% (33/162) |
| Balanced translocation, triploid | 0.4% (4/908)<sup>37</sup> | N. A | 2% (4/201)<sup>42</sup> | N. A | N. A | N. A |
| CMA level | 5.0% (19/382)<sup>6</sup> | 6.0% (19/317)<sup>6</sup> | 5.4% (38/699)<sup>6</sup> | 1.9% (2/104) | 4.0% (1/25) | 2.3% (3/129) |
| SNV | N. A | N. A | 22% (11/49)<sup>5</sup><br>4.3% (3/69)<sup>6</sup> | 11.8% (12/102) | 50.0% (12/24) | 19.0% (24/126) |
| Intragenic CNV (50bp-100kb) |  |  |  | 3.3% (3/90) | 0 | 2.9% (3/102) |

N.A: not available

SNVs we found 14% diagnostic rate in the whole cohort and 19% positive rate in cases chromosomal anomaly and pCNVs were not found. Earlier research by Fu et al^37^ which had 65 fetuses with CNS malformations reported a diagnostic yield of 23.1% and a 24% in 196 fetuses with all types of malformations. A latest prospective cohort study by Petrovski^5^ reported an over diagnostic rate of 10% of ES in fetuses having structural anomaly prenatally with 243 parent-fetus trios, and the diagnostic genetic variants in CNS subgroup was 22%(11/49). Another large prenatal exome sequencing study including 610 nuclear families by Lord^6^ published simultaneously with Petrovski’s reported an overall diagnostic yield of 8.5% and 3% in 69 fetuses with brain anomaly.

The three studies followed similar clinical procedure of ultrasound evaluation, karyotyping and CMA, and then ES, while the SNV diagnosis yield varied a lot among studies. This may be due to different sample size, distribution of fetal phenotypes and the executed criterion of variants classification. Moreover, with 162 patients, our study is currently the largest one focusing on fetal sonographic CNS anomalies. Although the majority of samples was collected after termination or spontaneous fetal death, we included patients in an unselective way in which conditions that have less associations with monogenic basis such as NTDs, isolated arachnoid cyst and even destructive cerebral lesions were included because we aimed at demonstrating the genomics bases of common fetal CNS anomalies rather than promoting diagnosis rate of genetic tools by carefully selecting patients.

It is worth mentioning that our GS approach can identify intragenic CNVs beyond the detecting limit of currently CMA. In this study, we systematically evaluated their possible causal relationship to fetal anomalies. Although there are several genetic testing panels designed for special diseases may involve probes or primers for intragenic CNVs, they have not been routinely investigated across a wide range of disease genes in traditional genetic testing. Their application scope is even more limited in prenatal as fetal abnormalities are usually difficult to get a definite diagnosis prenatally and the genetic background of fetal structural anomalies is highly heterogeneous. However, intragenic CNVs may contribute substantial part of pathogenic variants. Truty et al^39^ investigated the prevalence and properties of intragenic CNVs in >143,000 individuals referred for genetic testing, and found that ∼10% prevalence of intragenic CNVs among individuals with a positive test result and highest frequencies in neurological diseases. In pediatric and rare disorders, NF1 was the gene most frequently affected by pathogenic CNVs. In our study, intragenic CNVs analysis identified three likely pathogenic intragenic CNVs, bringing an extra 2.9% of diagnosis. In fact, we did find other intragenic CNVs encompassing internal exons and predicted to have an adverse effect on the transcript reading frame on genes with loss-of-function (LOF) mutational mechanisms. However, there isn’t solid evidence supporting their association with fetal CNS anomalies. Therefore, we believed that if intragenic CNV could be more widely analyzed in research or clinical laboratories and prenatal imaging manifestations be more widely available in public databases like Decipher or ClinVar, the diagnostic yield of intragenic CNVs would be largely improved.

There are concerns that GS requests more fetal DNA and longer turnaround time and higher cost^6^. While this is not true. The DNA needed is depended on the protocols of library construction instead of on sequencing itself. 1μg of genomic DNA is usually used as the standard protocol across popular platforms ^40^. TruSeq Exome Library Prep protocol (November 2015) used 100ng starting DNA with Covaris fragmentation. MGI Easy Universal DNA Library Prep Set from MGI starting from 0.5-50ng is available for the platforms we applied when fetal DNA is extremely limited. Moreover, GS need extra DNA for CMA as only a single GS sequencing is needed. Therefore, DNA amount is not a problem. As for turnaround time, according to previous studies, it varies from 2-15 weeks to obtain and interpret ES results routinely ^5^. Our GS routine turnaround time is 2-3 weeks including chromosomal anomalies, CNVs, SNVs and intragenic CNV interpretation per sample and another week for sanger validation. In fact, it has been reported in two previous studies that rapid GS takes only 26-50 hours for emergency management of genetic diseases or in neonatal intensive care unit ^16,41^. GS is as rapid as ES in a routine protocol and even faster if taking account of extra time for CMA as ES’s prerequisite. Besides, the sequencing cost of one GS is lower than CMA plus ES and other cost is similar to ES. In fact, though we sequenced samples to ∼40x depth, variant identification performance is acceptable when depth decreased to 20-30x, which will further cut down the cost of GS.

Human Phenotype Ontology (HPO) database is useful to select candidate genes^9^. However, we should keep it in mind that ultrasound findings in the brain could be unspecific and developing, and minor anomalies could be missed or undiagnosed prenatally. For example, though signs of abnormal cortical development are possible to be discovered by seasoned experts through advanced neurosonography and MRI, it is difficult to be confirmed in mid-semester. While with the help of GS, widen of lateral ventricles and subarachnoid space as in P431 and P796 were confirmed to be lissencephaly. Therefore, for enhancing a genetic diagnosis of fetal CNS anomalies, to describe abnormal ultrasonography manifestations may be more helpful than to give a disease diagnosis.

While GS is a suitable testing modality for fetus CNS anomalies, our approach has some limitations. Uniparental disomy and triploidy were not analyzed in this study since such pipelines needed further development. And it is now difficult to determine the clinical significance of variants called in intron and noncoding area due to lacking of large scale databases. Even though we did find some variants in intron that were previously reported to be likely pathogenic, but we considered them more likely to be benign due to high frequency in our control database. Continuing to this pilot study which concerned CNS anomalies, more cases of fetal ultrasound abnormalities of each anatomic system are being studied now, and the potential power of GS will be further demonstrated.

## Data Availability

https://db.cngb.org/cnsa/

## Acknowledgement

This project is mainly supported internally by BGI-Shenzhen. It is also supported by Science, Technology and Innovation Commission of Shenzhen Municipality under grant No. JCYJ20180703093402288, Department of Science and Technology of Guangdong Province under grants No. 2019B020227001, Natural Science Foundation of Hubei Province (2017CFB789) and Innovation Team Project of Health commission of Hubei Province (WJ2018H0132). Sequencing data was produced by China National Genebank. We are also grateful to BGI-Wuhan for helping transferring samples.

